# Trends and Future Projections of Liver Cirrhosis Burden in Sub-Saharan Africa with Hepatitis B Vaccination Impact

**DOI:** 10.1101/2025.08.19.25333950

**Authors:** Ellis K. Paintsil, Kai Yuan, Ruqayyah Labidi, Debbie L. Shawcross

**Affiliations:** Roger Williams Institute of Liver Studies, School of Immunology and Microbial Sciences, Faculty of Life Sciences and Medicine, King’s College London, UK; School of Bioscience Education, Faculty of Life Sciences and Medicine, King’s College London, UK; School of Global Affairs, Faculty of Social Science & Public Policy, King’s College London, UK

**Keywords:** Liver cirrhosis, Chronic liver disease, Global Burden of Disease, Sub-Saharan Africa, Hepatitis B vaccination

## Abstract

Liver cirrhosis and chronic liver diseases impose a substantial and growing health burden globally, with sub-Saharan Africa (SSA) disproportionately affected. Leveraging Global Burden of Disease 2021 data and Bayesian hierarchical models, we quantified mortality and disability trends in SSA from 1990 to 2021 and projected disease burden through 2035, incorporating scenarios for hepatitis B vaccination scale-up.

In 2021, liver cirrhosis accounted for an estimated 181,311 deaths in SSA, despite a 29% decline in age-standardized death rates (ASDR) since 1990. Absolute deaths increased by 65%, predominantly driven by hepatitis B (37%), hepatitis C (28%), and alcohol-related cirrhosis (17%). Disability-adjusted life years (DALYs) surged by 76%, from approximately 1.05 million in 1990 to 1.85 million in 2021, highlighting rising absolute disability alongside a 29% reduction in age-standardized DALYs. Mortality and disability burdens were highest in Somalia, Central African Republic, and Guinea-Bissau. Males bore nearly twice the burden of females. While death rates declined across all socio-demographic strata, absolute deaths rose by 55–86%. Projections to 2035 suggest further potential reductions in mortality from hepatitis B (up to 21.5%), hepatitis C (up to 18.7%), and alcohol-related cirrhosis, while the burden of non-alcoholic fatty liver disease is expected to remain stable or increase slightly. Scaling up hepatitis B vaccination could further avert 27% of related deaths by 2035.

These findings reveal persistent and widening disparities in cirrhosis burden across SSA, underscoring the urgent need for integrated, context-specific interventions combining viral hepatitis control with metabolic liver disease management to improve equitable liver health outcomes.

## Background

Liver cirrhosis, other chronic liver diseases, and liver cancer are among the leading global causes of premature mortality and disability, responsible for two million deaths annually (Devarbhavi et al., 2023; Gan et al., 2025). In Africa, the burden has been reported to be disproportionately high and is expected to continues to rise (Spearman, 2023; Wu et al., 2024; Younossi et al., 2023) amid rapid population growth, demographic transitions, and fragile health systems (Gouda et al., 2019; Spearman & Sonderup, 2015). Despite progress in the global prevention and management of liver disease, sub-Saharan Africa (SSA) has lagged behind due to limited access to hepatitis B virus (HBV) vaccination, diagnostic services, and affordable treatment (Kramvis, 2020; Spearman et al., 2023). Adding to these challenges is the growing threat of antimicrobial resistance (AMR), with regionally variable but high rates of multidrug-resistant reported among cirrhosis patients (Paintsil et al., 2025), underscoring the urgent need for targeted infection control and surveillance. The region’s young and expanding population, coupled with health infrastructure constraints and competing health priorities, has created a challenging environment for liver disease control (Grinin & Korotayev, 2023; Spearman & Sonderup, 2015). Yet, SSA remains significantly underrepresented in global liver disease surveillance and policy discussions, limiting the region’s ability to mount effective, context-specific responses.

The causes and distribution of liver cirrhosis in SSA are thought to be highly heterogeneous, shaped by a complex interplay of infectious and non-communicable drivers (Boudreaux et al., 2020; Lan et al., 2023; Spearman & Sonderup, 2015). Chronic viral hepatitis particularly HBV and HCV remain endemic in many settings and is suspected to contribute significantly to the burden (Spearman et al., 2023). Meanwhile, alcohol-related liver disease is emerging as a major concern, with rising harmful consumption patterns across many SSA countries (Ferreira-Borges et al., 2017; Huang et al., 2023). Non-alcoholic fatty liver disease (NAFLD), long under-recognized, may also be gaining relevance due to urbanization and increasing metabolic risk (Younossi et al., 2016). These shifting patterns are compounded by systemic inequities such as gender disparities, fragile health systems, and limited access to diagnosis and care but their role in shaping liver disease outcomes in SSA remains poorly quantified (Spearman & Sonderup, 2015).

Despite the growing importance of cirrhosis and chronic liver diseases in SSA, comprehensive, regionally disaggregated data remain scarce. Prior global assessments have often aggregated SSA into a single unit or excluded detailed analysis, masking important sub-regional patterns and trends (Guo et al., 2025; Lan et al., 2023, 2023; Tham et al., 2025; Wu et al., 2024). To address this gap, we conducted a granular analysis of liver cirrhosis and chronic liver disease burden across SSA using Global Burden of Disease (GBD) 2021 estimates. This study provides stratified insights by sex, aetiology, country, and socio-demographic index (SDI), and models future burden through 2035, including a vaccination scale-up scenario. By illuminating disparities and epidemiological shifts, our findings aim to inform regionally tailored strategies that address both infectious and non-infectious liver disease threats in SSA—contributing to efforts to reduce avoidable liver-related mortality and disability across the region.

## Methods

### Data Source and Variables Extracted

This study used publicly available estimates from the GBD 2021 study (Institute for Health Metrics and Evaluation [IHME], 2024). Age-standardised mortality and disability-adjusted life years (DALYs) attributable to liver cirrhosis and other chronic liver diseases in SSA were extracted for the period 1990–2021 from the IHME GBD Results Tool (https://vizhub.healthdata.org/gbd-results).

We extracted data on the following parameters: measure (deaths and DALYs), metric (rate and number), and cause—including NAFLD, cirrhosis and other chronic liver diseases, chronic hepatitis B (including cirrhosis), chronic hepatitis C (including cirrhosis), alcohol-related cirrhosis, and cirrhosis due to other causes. Estimates were disaggregated by sex (male, female, both), age group (all ages and age-standardised), year (1990–2021), and geography—including SSA overall, its four subregions (Western, Central, Eastern, and Southern), and 46 individual countries and territories (Supplementary Tables S1-S3). The SDI for each country was also extracted and used to classify countries into SDI quintiles: low, low-middle, and middle (Supplementary File 1).

### Projection and Intervention Scenario Modelling

To project the burden of liver cirrhosis in SSA through 2035, we developed a Bayesian hierarchical time-series model using Integrated Nested Laplace Approximation (INLA) (Rue et al., 2009). The model was trained on GBD estimates from 1990 to 2021 and accounted for covariates including age, sex, aetiology, and SDI. We applied a second-order random walk (RW2) prior to capture smooth temporal trends in log-transformed age-standardised death rates (ASDRs) and DALYs (Blangiardo & Cameletti, 2015; Rue et al., 2017). Projections were generated under a baseline scenario assuming the continuation of historical trends, with 95% credible intervals reflecting both parameter uncertainty and data variability.

In addition, an intervention scenario was modelled to estimate the potential impact of scaling up hepatitis B vaccination starting in 2026, in line with WHO elimination targets (WHO, 2021). This scenario assumed an increase in hepatitis B birth-dose (HepB-BD) coverage from approximately 17% in 2021 to 50% by 2030, and an increase in three-dose infant vaccination (HepB3) coverage from around 75% to 90% by 2030, sustained thereafter (Kabore, 2023). Based on prior dynamic transmission modelling studies and assuming 90% vaccine efficacy against chronic HBV infection (de Villiers et al., 2021; Nayagam et al., 2023), we applied a linear reduction of 25% in HBV-related cirrhosis ASDRs and 30% in related DALYs from 2026 to 2035. These reductions were applied directly to the baseline projections to reflect the anticipated long-term population-level impact of improved vaccination coverage. The intervention effect was implemented as a simplified adjustment to the baseline projections and does not explicitly model HBV transmission dynamics.

### Statistical Analysis

Trends in total deaths and ASDRs from 1990 to 2021 were described using summary statistics, with 95% uncertainty intervals (UIs) to reflect GBD-derived uncertainty. Average Annual Percent Change (AAPC) in ASDR and ASDALYs was estimated using log-linear regression models fitted to the log-transformed rates over time (Clegg et al., 2009; Kim et al., 2000). Pooled estimates of ASDR and ASDALYs across subregions were calculated using inverse variance–weighted meta-analysis, with 95% confidence intervals derived from standard errors calculated from reported uncertainty intervals. Bayesian projections of ASDRs and DALYs through 2035 were performed using INLA with RW2 priors as noted above (Rue et al., 2009), and included both baseline and intervention scenarios. Linear reductions were also used for sensitivity analysis of intervention effects. We assessed the ecological relationship between country-level SDI and ASDR/ASDALYs using Spearman rank correlation (Zar, 1972). All statistical analyses and visualisations were conducted in R (version 4.5.1), using the INLA, ggplot2, and other relevant packages for regression, meta-analysis, and data wrangling (Lindgren & Rue, 2015; R Core Team, 2024; Wickham, 2016)

## Results

### Burden and Trends of Liver Cirrhosis and Chronic Liver Diseases in SSA, 1990–2021

In 2021, liver cirrhosis and other chronic liver diseases caused an estimated 181,311 deaths (95% UI: 155,374–208,123) in SSA, corresponding to an ASDR of 33.2 per 100,000 population (95% UI: 29.2–37.7). Although the ASDR declined by 29% since 1990, when it was 46.6 (95% UI: 40.8–55.9) per 100,000, total deaths increased by 65%, from 109,694 (95% UI: 96,718–129,521) in 1990. Chronic hepatitis B-related cirrhosis accounted for the largest proportion of deaths in 2021 (37%, n = 67,434), followed by chronic hepatitis C (28%, n = 51,344) and alcohol-related cirrhosis (17%, n = 30,647). Temporal trends in total and cause-specific cirrhosis deaths are shown in Supplementary Figure 1A–F. Regionally, Central SSA had the highest ASDR in 2021 at 40.3 per 100,000 population (95% UI: 30.3–51.2), followed by Eastern (38.5; 95% UI: 33.9–43.2), Western (31.0; 95% UI: 25.1–36.4), and Southern SSA (17.8; 95% UI: 15.5–20.3) (Figure 1A–B; Table 1). Eastern SSA recorded the highest absolute number of deaths, 72,973 (95% UI: 63,805–82,342), followed by Western, Central, and Southern SSA (Table 1). Mortality rates were consistently higher in males than females (Table 1). In 1990, the ASDR in males was 61.1 per 100,000 (95% UI: 51.8–76.6), decreasing to 44.6 (95% UI: 37.8–51.2) in 2021. Corresponding rates in females were 32.2 (95% UI: 26.3–38.3) in 1990 and 23.1 (95% UI: 20.3–26.8) in 2021, reflecting a male-to-female ratio of approximately 1.9:1 in 2021 (Figure 1C–D; Table 1). This disparity was even greater in alcohol-related cirrhosis, with males experiencing an ASDR of 9.9 (95% UI: 7.2–13.1) compared to 2.6 (95% UI: 1.9–3.4) in females, representing a 3.8-fold difference. Similarly, for hepatitis B-related cirrhosis, the ASDR was 17.1 (95% UI: 13.8–20.8) in males and 8.5 (95% UI: 6.8–10.3) in females (Figure 1E–F).

**Figure 1.**
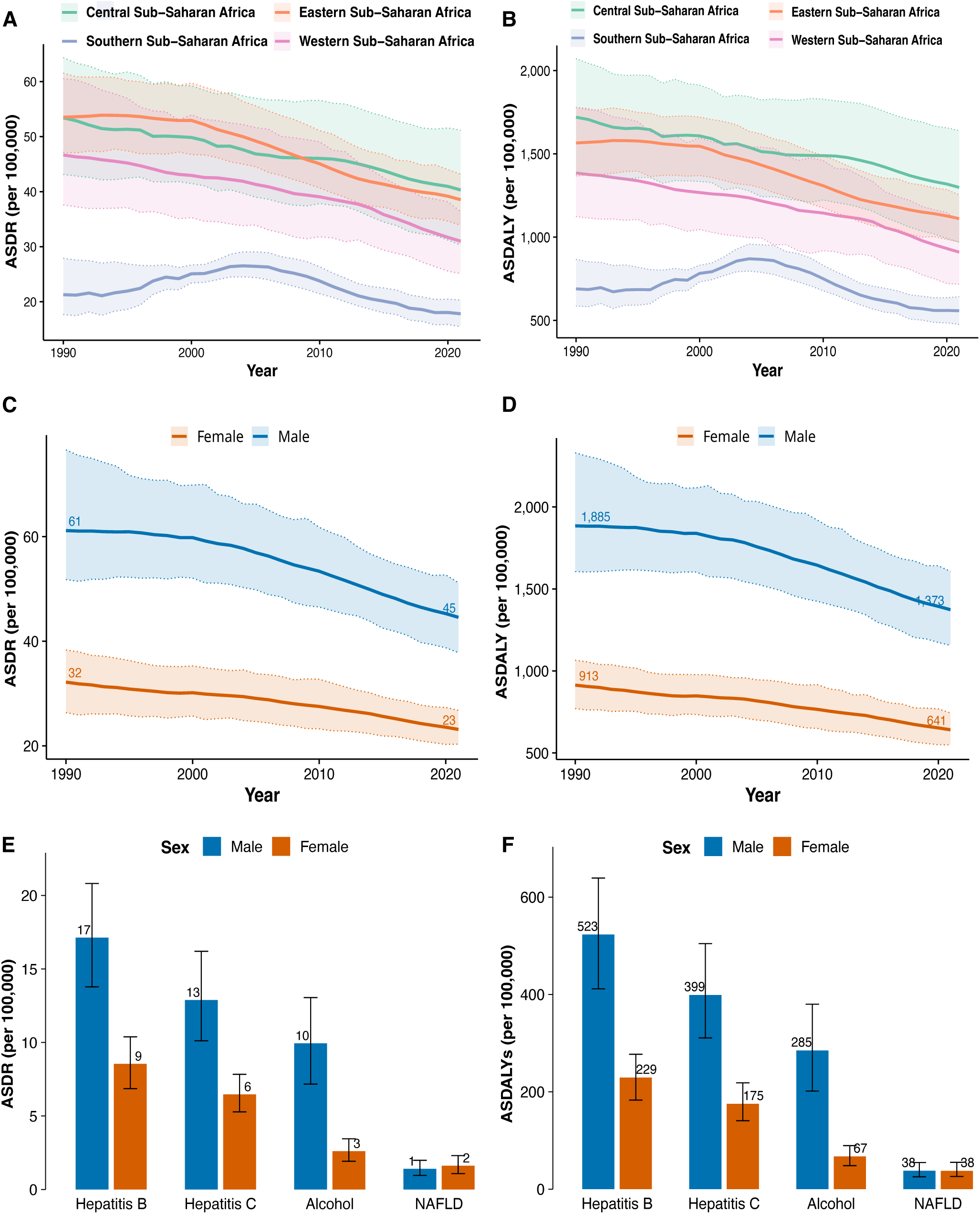
Trends in Age-standardised death rates (ASDR) and disability-adjusted life years (ASDALYs) for cirrhosis and other chronic liver diseases by sub-region, sex, and aetiology in Sub-Saharan Africa. ASDR trends by geographic sub-region from 1990 to 2021 **(A)** and ASDALY trends by sub-region over the same period **(B)**. Temporal trends in ASDR **(C)** and ASDALYs **(D)** stratified by sex, with shaded ribbons representing 95% uncertainty intervals. Sex-stratified ASDRs **(E)** and ASDALYs **(F)** for major cirrhosis aetiologies—hepatitis B, hepatitis C, alcohol use, and non-alcoholic fatty liver disease—in 2021. Bars show point estimates with error bars indicating 95% uncertainty intervals.

**Table 1.**
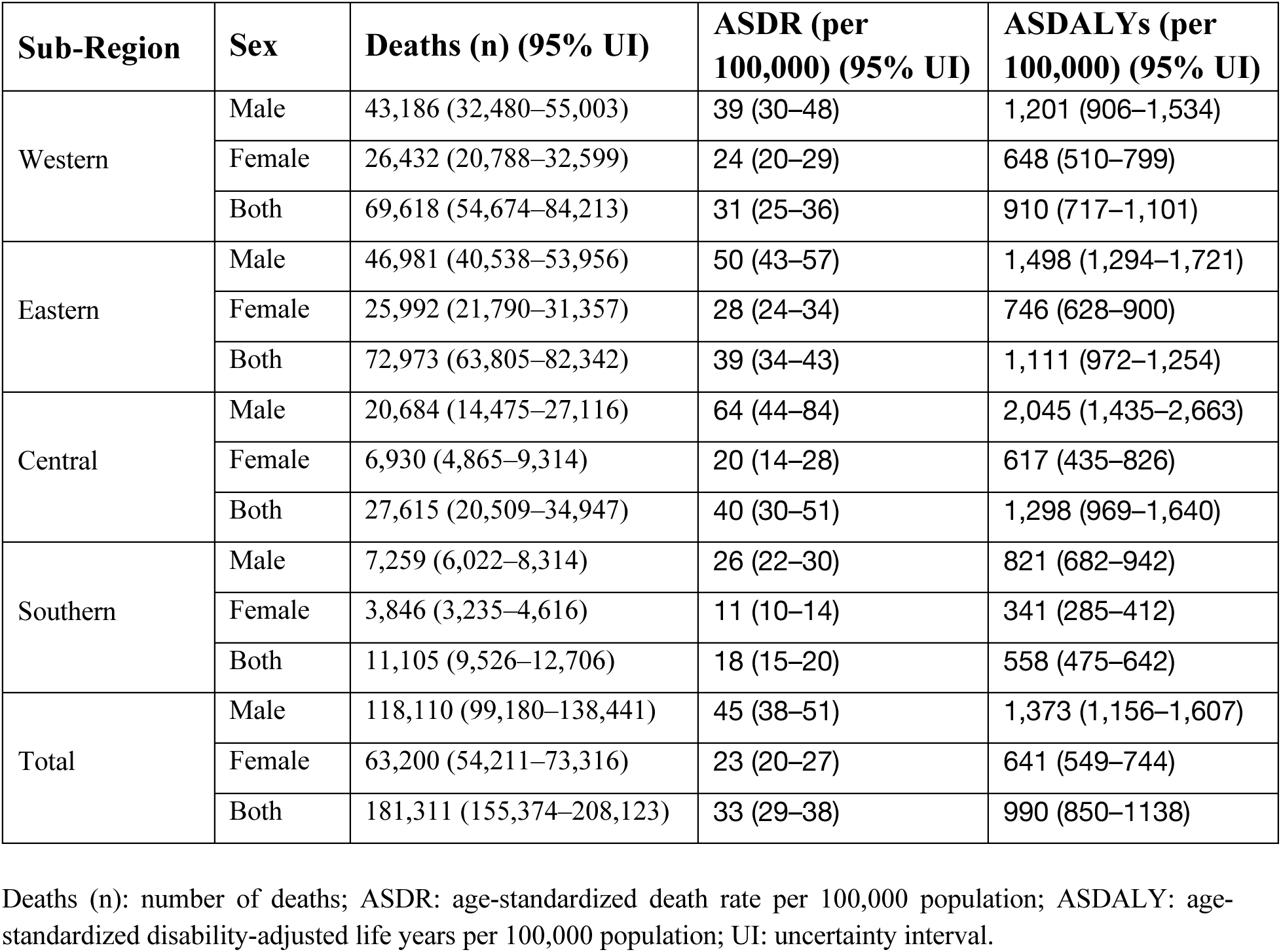
Burden of liver cirrhosis and other chronic liver diseases in sub-Saharan Africa by subregion and sex in 2021.

### Geographic and Sex-Specific Variations in Cirrhosis Mortality and Disability-Adjusted Life Years

The DALY burden attributable to liver cirrhosis and other chronic liver diseases increased by 76%, rising from 1,049,815 (95% UI: 839,461–1,318,915) in 1990 to 1,851,307 (95% UI: 1,433,585–2,298,023) in 2021. However, after adjusting for population growth and ageing, the ASDALY declined by 29%, from 1,400 (95% UI: 1,236–1,652) to 990 (95% UI: 850–1,138) per 100,000 population over the same period. NAFLD contributed the smallest share of the 2021 burden at 38 per 100,000 (95% UI: 26–54), while hepatitis B and hepatitis C were the leading drivers, accounting for 369 (95% UI: 297–445) and 282 (95% UI: 223–351) per 100,000, respectively. Central SSA recorded the highest ASDALY at 1,319 per 100,000 (95% UI: 992–1,656), followed by Eastern (1,111; 95% UI: 972–1,254) and Western (910; 95% UI: 717–1,101) (Figure 2A-B). The ASDALY burden was substantially higher in males, at 1,373 per 100,000 (95% UI: 1,156–1,607), compared to females, at 640 per 100,000 (95% UI: 549– 744), representing an approximate 2.1-fold excess.

**Figure 2.**
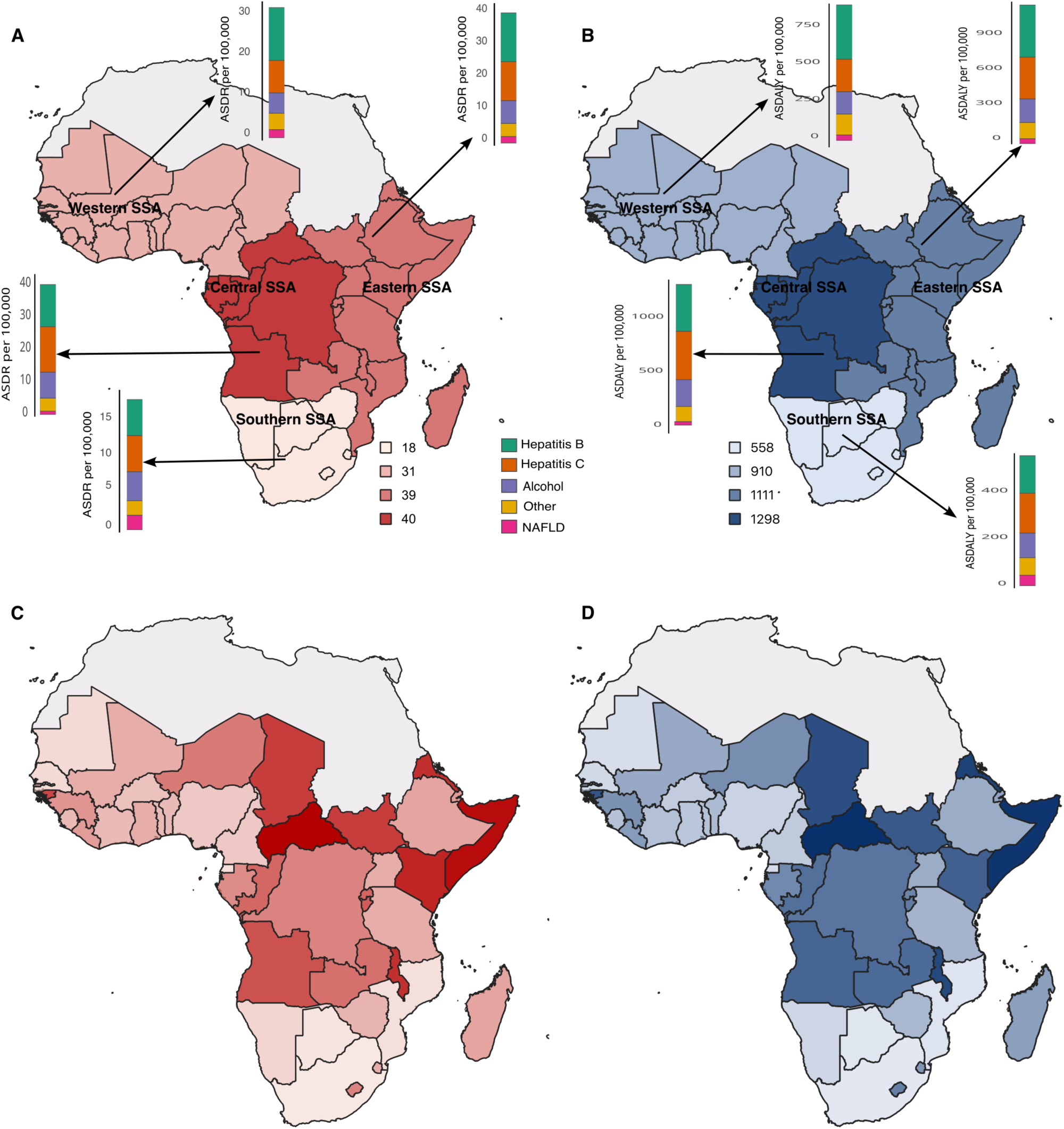
ASDR and ASDALYs in for cirrhosis aetiologies and burden across regions and countries in SSA, 2021. ASDR **(A)** and ASDALYs **(B)** by sub-region. Bars for each sub-region represent AAPC estimates by cirrhosis aetiology—hepatitis B, hepatitis C, alcohol use, non-alcoholic fatty liver disease, and other causes. ASDR **(C)** and ASDALYs **(D)** at the country level across SSA in 2021. Deeper red or blue colours indicate more negative AAPCs, reflecting faster declines in cirrhosis burden.

In 2021, the highest ASDRs from liver cirrhosis and other chronic liver diseases among SSA countries were recorded in the Central African Republic (63 per 100,000; 95% UI: 48–81), Somalia (59; 95% UI: 40–83), and São Tomé and Príncipe (53; 95% UI: 33–71) (Figure 2C). Corresponding ASDALYs were also elevated in these countries, with the Central African Republic recording the highest burden at 2,089 per 100,000 (95% UI: 1,574–2,747), followed by Somalia (1,770; 95% UI: 1,166–2,497) and São Tomé and Príncipe (1,496; 95% UI: 942– 2,082) (Figure 2D). In contrast, the lowest ASDRs were observed in South Africa (15; 95% UI: 14–16), Botswana (18; 95% UI: 13–22), and Mozambique (24; 95% UI: 17–31), with correspondingly lower ASDALYs of 462 (95% UI: 421–509), 528 (95% UI: 408–671), and 674 (95% UI: 492–878) per 100,000, respectively (Figure 2C-D).

### Burden of Cirrhosis and Chronic Liver Diseases by Aetiology Across Sub-Saharan African Countries

The burden of cirrhosis and other chronic liver diseases in SSA varies substantially by country and underlying aetiology. Chronic hepatitis B, including cirrhosis, has been the leading contributor to both ASDR and ASDALYs in most countries between 1990 and 2021 (Supplementary Figure 3). In 2021, the highest ASDALY rates for hepatitis B-related cirrhosis were observed in Somalia (739; 95% UI: 449–1090), Central African Republic (711; 95% UI: 454–997), and Guinea-Bissau (617; 95% UI: 392–879). Chronic hepatitis C-related cirrhosis also contributed substantially, particularly in Central African Republic (717; 95% UI: 489– 1017), Eritrea (471; 95% UI: 311–662), and Angola (459; 95% UI: 325–636). Alcohol-related cirrhosis exhibited marked geographic heterogeneity, with the highest ASDALYs in Central African Republic (384; 95% UI: 231–598) and Somalia (311; 95% UI: 177–506). NAFLD, including cirrhosis, was associated with the lowest ASDALY rates overall but was more prominent in the Kingdom of Eswatini (83; 95% UI: 50–134) and São Tomé and Príncipe (72; 95% UI: 39–113).

### Burden of Liver Cirrhosis by Socio-demographic Index in Sub-Saharan Africa

The burden of cirrhosis declined with increasing SDI: low-middle SDI countries had an ASDR of 31.5 (UI: 28.7–34.2) and ASDALY of 920 (UI: 837–1,003), while middle SDI countries had substantially lower estimates (ASDR: 15.9; UI: 14.1–16.6; ASDALYs: 480; UI: 439–521). This inverse gradient was confirmed in correlation analyses, with SDI negatively associated with both ASDR (Spearman ρ = –0.35, *p* = 0.016) and ASDALY (ρ = –0.34, *p* < 0.02). Countries with the lowest SDI, such as Somalia and the Central African Republic, experienced disproportionately high cirrhosis-related mortality and disability burdens (Figure 3A-B). HBV was the leading cause of cirrhosis-related mortality in low SDI countries, accounting for 39% of deaths (13.5; UI: 12.6–14.5), compared to 29% (4.4; UI: 3.8–5.0) in middle SDI settings. The contribution of NAFLD increased with SDI, rising from 3% of deaths in low SDI countries (NAFLD-specific ASDR: 32; UI: 29–35) to 9% in middle SDI countries (27; UI: 20–34). HCV accounted for 31% of cirrhosis deaths in both low (9; UI: 9–10) and middle SDI countries (5; UI: 4–5), but declined to 17% in low-middle SDI settings (5; UI: 5–6). Across all SDI strata, cirrhosis mortality was consistently higher in males. The male-to-female ASDR ratio ranged from 1.9 in low SDI countries to 2.2 in middle SDI countries.

**Figure 3.**
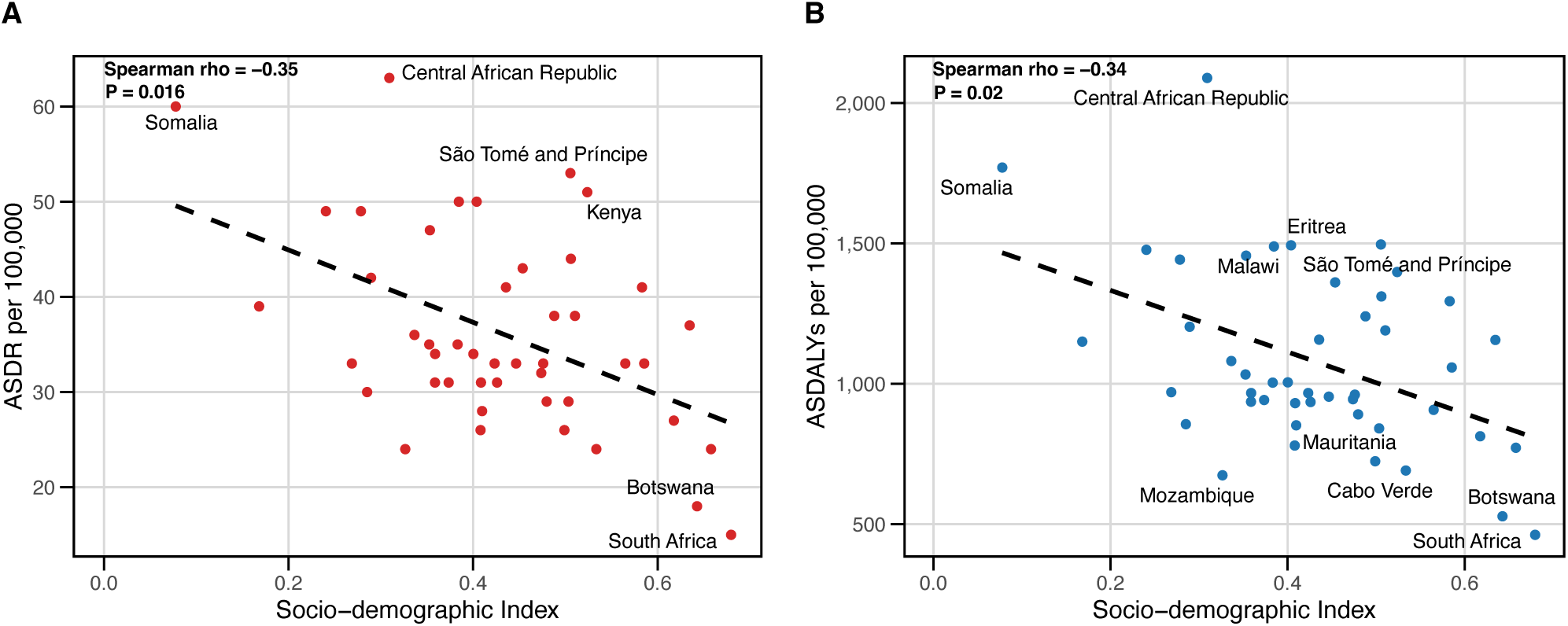
Association between SDI and the burden of cirrhosis in SSA, 2021. ASDR per 100,000 population plotted against SDI for each country **(A)**. ASDALYs per 100,000 population plotted against SDI **(B)**. Each point represents a country, with dashed lines indicating linear regression fits. Countries with extreme values (outliers in ASDR or ASDALY) are labelled. Spearman correlation coefficients and p-values are shown.

Temporal trends from 1990 to 2021 revealed significant declines in ASDRs across all SDI strata. The greatest AAPC was observed in middle SDI countries (−1.96%, 95% CI: −2.12 to −1.80), followed by low SDI (−0.97%, 95% CI: −1.09 to −0.89) and low-middle SDI settings (−0.78%, 95% CI: −0.94 to −0.98). At the country and regional levels, AAPC estimates for ASDR are presented in Supplementary Table S4, and those for ASDALY are presented in Supplementary Table S5. Despite these reductions, the absolute number of cirrhosis-related deaths increased by approximately 86% in low SDI countries, 79% in low-middle SDI countries, and 55% in middle SDI countries over the study period. These patterns were consistent across all sub-regions and aetiologies, as shown in Supplementary Figure 2, which presents AAPC estimates for ASDRs and ASDALYs by sub-region stratified by cirrhosis aetiology (A–B), along with country-level estimates (C–D).

### Projected Burden of Liver Cirrhosis and Chronic Liver Diseases in SSA, 2022–2035

Using a Bayesian hierarchical model trained on observed trends from 1990 to 2021, we projected ASDRs and ASDALYs for liver cirrhosis and chronic liver diseases across SSA sub-regions from 2022 to 2035 (Supplementary Table 6 and 7).

Projections reveal varied regional patterns: in Western SSA (Figure 4A), hepatitis B-related ASDRs are expected to decline by 13.3% (12.8 to 11.1 per 100,000), hepatitis C by 11.8% (7.6 to 6.7), and alcohol-related cirrhosis by 15.1% (5.3 to 4.5), while NAFLD-related ASDRs slightly increase by 6.7% (1.5 to 1.6). Eastern SSA (Figure 4B) follows a similar pattern with hepatitis B decreasing 15.9% (14.5 to 12.2), hepatitis C by 12.9% (11.6 to 10.1), alcohol-related deaths by 11.8% (6.8 to 6.0), and stable NAFLD rates at 1.6 per 100,000. Central SSA (Figure 4C) exhibits the largest absolute declines: hepatitis B drops 21.5% (13.5 to 10.6), hepatitis C 18.7% (13.9 to 11.3), and alcohol-related cirrhosis 14.8% (8.1 to 6.9), while NAFLD-related ASDRs rise sharply by 25% (1.2 to 1.5). Southern SSA (Figure 4D), with the lowest cirrhosis burden, shows modest decreases in hepatitis B (7.8%; 5.1 to 4.7) and hepatitis C (5.6%; 5.4 to 5.1), stable alcohol-related mortality at 3.6 per 100,000, and a slight 5.9% decline in NAFLD (1.7 to 1.6). Aggregated projections across total SSA (Figure 4E) suggest a modest decline in ASDRs and ASDALYs for most cirrhosis aetiologies, whereas NAFLD-related burden remains relatively stable in almost all subregions (Figure 4A–E; Supplementary Figure 3A–E).

**Figure 4.**
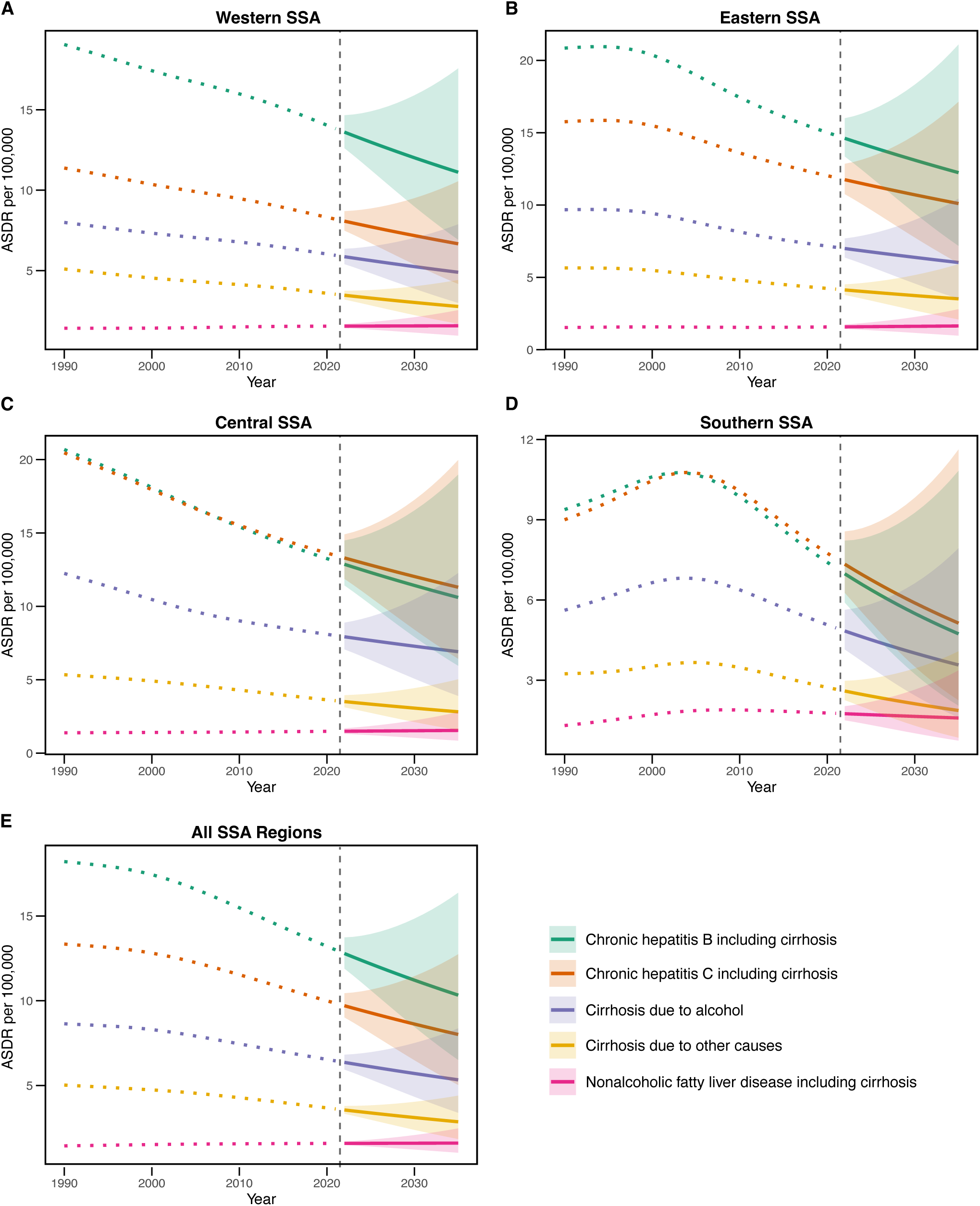
Projected ASDR for cirrhosis aetiologies across SSA, 2022–2035. Panels **(A)–(E)** show ASDR projections per 100,000 population for Western SSA, Eastern SSA, Central SSA, Southern SSA, and total SSA, respectively. Coloured dotted lines represent observed data for each aetiology from 1990 to 2021, while solid lines indicate corresponding projections from 2022 to 2035. Shaded ribbons denote 95% uncertainty intervals around the projections. The vertical dashed line marks the boundary between observed and projected periods.

Sex-stratified projections show that males will continue to experience a disproportionately higher burden of cirrhosis-related morbidity and mortality across all aetiologies. Between 2021 and 2035, ASDRs are projected to decline from 44.6 to 38.8 per 100,000 in males and from 23.1 to 18.9 in females. Correspondingly, ASDALYs are expected to fall from 1,373 to 1,175 per 100,000 in males and from 641 to 522 in females. The male-to-female burden ratio is projected to remain close to 2:1 throughout the projection period (Supplementary Figure 4A-B).

Figure 5 highlights regional disparities in projected cirrhosis burden across SSA. Central SSA is projected to maintain the highest ASDRs and ASDALYs through 2035, followed by Eastern, Western, and Southern SSA.

**Figure 5.**
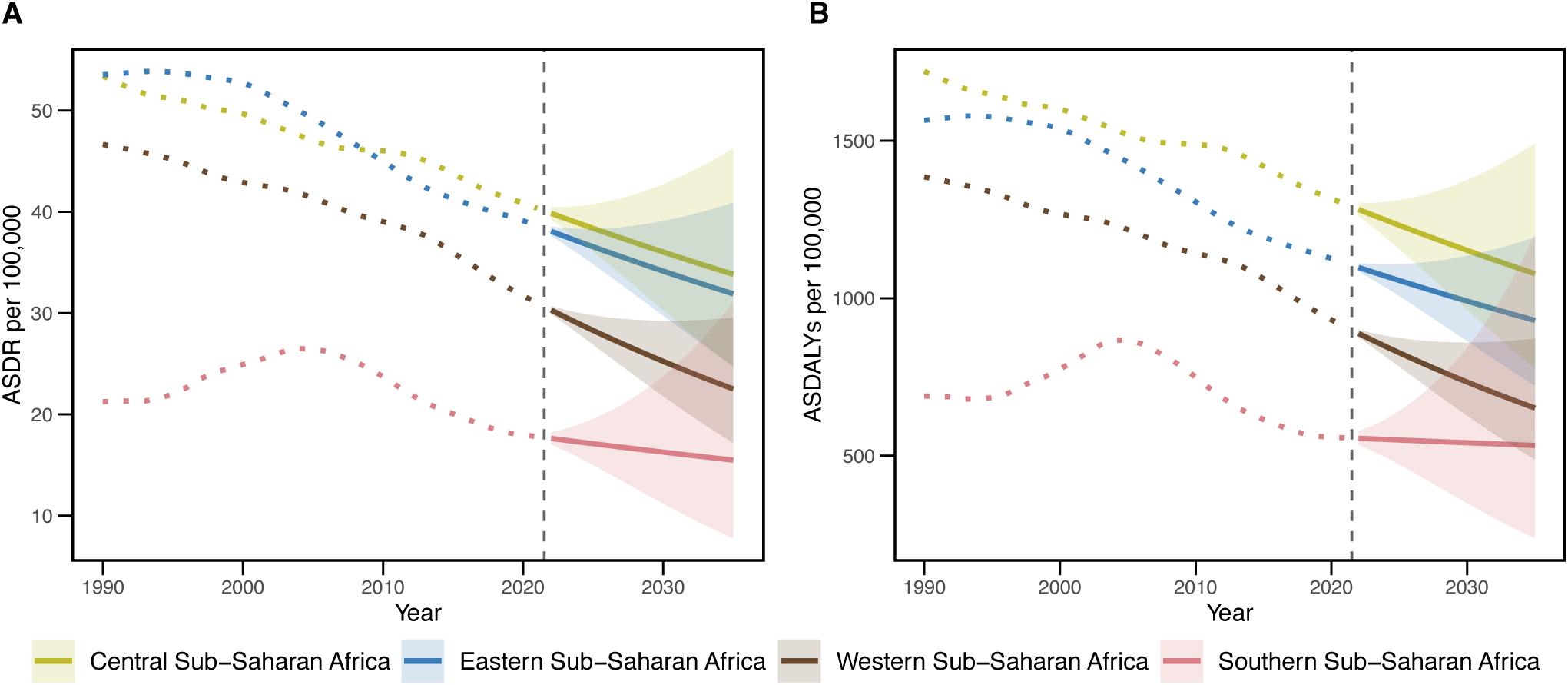
Projected ASDR and ASDALYs for cirrhosis and other chronic liver diseases across SSA sub-regions, 2022–2035. Panels show ASDRs **(A)** and ASDALYs **(B)** per 100,000 population by sub-regions. Coloured dotted lines represent observed data (1990–2021) and solid lines indicate projections from 2022 to 2035. Shaded ribbons denote 95% uncertainty intervals. The vertical dashed line separates observed and projected periods.

### Projected Reduction in HBV-Related Cirrhosis Burden Following Vaccination Scale-Up

Under the hepatitis B vaccination scale-up scenario, the projected ASDR for HBV-related cirrhosis in SSA declines from 8.4 to 6.1 per 100,000 population by 2035, representing a 27% reduction relative to baseline projections without intervention (Figure 6A). The corresponding ASDALYs decrease from 410 to 285 per 100,000, a 30% reduction (Figure 6B). Sensitivity analyses using a 20–35% effect range produced consistent directional results (Supplementary Tables S8 and S9).

**Figure 6.**
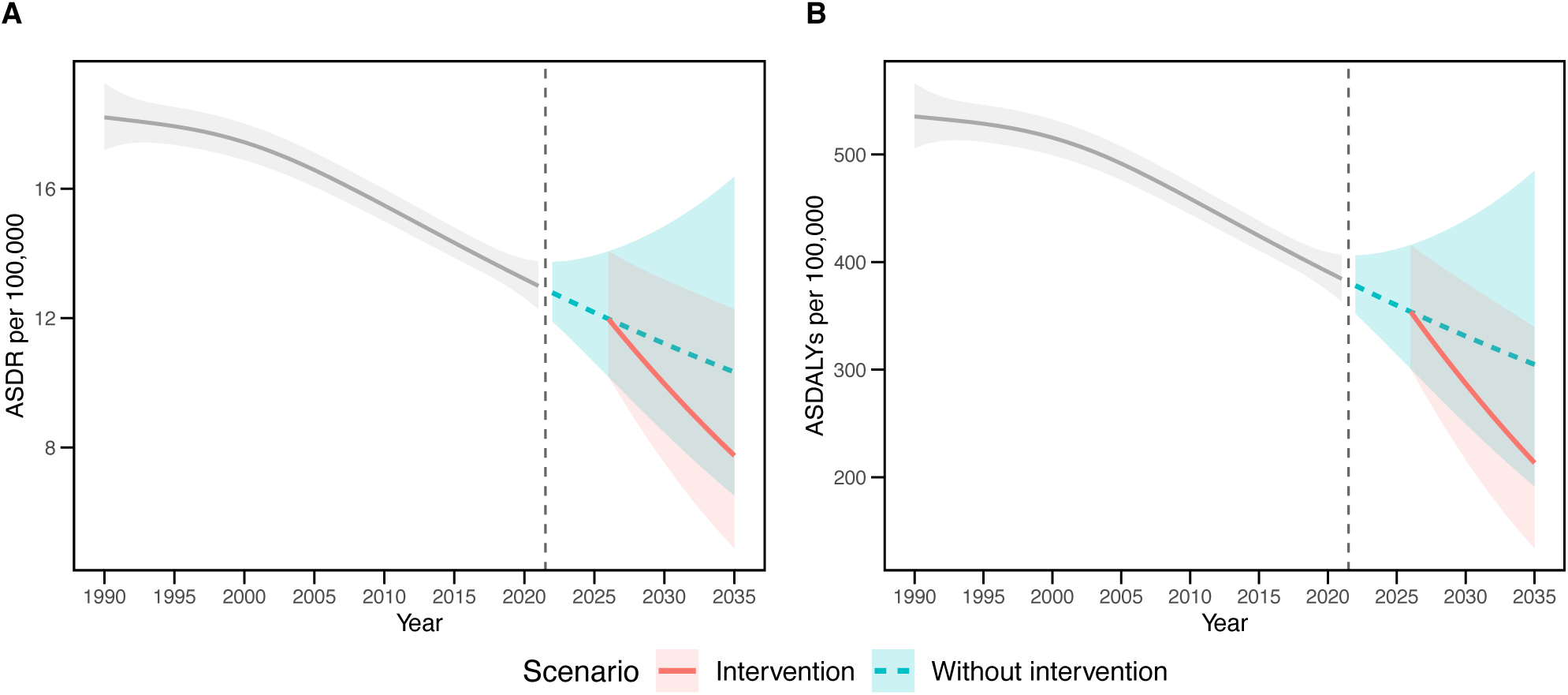
Projected impact of a hepatitis B vaccination intervention scenario on HBV-related cirrhosis burden in SSA, 2026–2035. Modelled trends in ASDALYs **(A)** and ASDR **(B)** per 100,000 population due to chronic hepatitis B, showing observed data from 1990–2021 followed by projections without intervention from 2022 and with a vaccination intervention scenario from 2026 onward. Shaded ribbons denote 95% uncertainty intervals. The vertical dashed line separates observed and projected periods.

## Discussion

This study provides the first comprehensive analysis of cirrhosis and chronic liver disease burden across SSA, incorporating age-standardised and absolute metrics disaggregated by sex, aetiology, and SDI. Our analysis shows that between 1990 and 2021, ASDRs and DALY rates declined by 29% and 26%, respectively. This decline likely results from incremental improvements in prevention, diagnosis and management within SSA, driven by wider use of affordable non-invasive diagnostics and accessible treatments such as non-selective beta blockers and hepatitis B and C interventions adapted to low-resource settings (Moed & Zaman, 2019; Sonderup et al., 2024). However, these declines have been offset by substantial increases in absolute numbers, with over 181,000 deaths and 1.85 million DALYs recorded in 2021, largely due to rapid population growth and ongoing demographic transition, with the region’s population projected to triple by 2100 (Ezeh et al., 2020). This divergence between rate reductions and rising crude burden aligns with trends observed in other low- and middle-income regions, where improvements in public health are often challenged by aging populations and expanding high-risk cohorts (Murray et al., 2018). Notably, the pronounced magnitude of these changes in SSA highlights persistent systemic challenges such as limited healthcare infrastructure, workforce shortages, and gaps in early diagnosis, treatment, and rise in AMR (Kruk et al., 2018; Paintsil et al., 2025), underscoring the urgent need for tailored health system strengthening and early intervention strategies to improve outcomes (Moghe et al., 2021).

Marked disparities in cirrhosis burden were evident across all SSA regions, with countries like the Central African Republic and Somalia consistently ranking among the highest in both mortality and DALYs. These patterns likely reflect the compounded effects of fragile health systems, political instability, and severely limited access to HBV vaccination, screening, and antiviral therapy in conflict-affected settings (Akingbola et al., 2025). In contrast, countries like South Africa and Botswana exhibited substantially lower burdens, possibly benefiting from relatively stronger health infrastructure. For example, Botswana has high geographic proximity to health facilities (Nkomazana, 2022) while South Africa benefits from better physician density and broader access to diagnostic and treatment services (Ahmat et al., 2022). Across all aetiologies, males experienced nearly double the burden compared to females —a disparity particularly pronounced in alcohol- and HBV-related cirrhosis. This may be driven by higher exposure to risk factors among men, including harmful alcohol consumption, late presentation to care, and lower engagement with preventive services (Boua et al., 2021; Yang et al., 2025; Yeatman et al., 2018). Socio-demographic development also shaped the burden; notably, middle SDI countries experienced the most pronounced declines in cirrhosis burden over time, suggesting that even modest improvements in socioeconomic conditions and health system capacity can yield meaningful reductions in disease burden (Tham et al., 2025). Strengthening equitable healthcare access and workforce capacity remains central to achieving sustained reductions in cirrhosis burden across the region.

Chronic viral hepatitis remains the predominant cause of cirrhosis across SSA, particularly in low-SDI countries, where HBV accounted for nearly 40% of cirrhosis-related deaths in 2021. While this aligns with HBV endemicity across the region (Sonderup & Spearman, 2024), the scale of its contribution underscores enduring gaps in prevention, screening, and treatment coverage (Spearman et al., 2023). HCV also plays a major role in liver-related mortality, especially in parts of Central and Eastern Africa, reflecting legacy issues around blood safety and healthcare-associated transmission (Karoney & Siika, 2013; Madhava et al., 2002). Meanwhile, alcohol-related cirrhosis displays marked spatial heterogeneity, often clustering in regions burdened by poverty, weak regulatory oversight, and limited harm reduction infrastructure (Manthey et al., 2019). Our analysis revealed that in Western and Eastern SSA, alcohol-related liver disease contributes disproportionately to the overall cirrhosis burden, driven by high per capita alcohol consumption and the absence of effective alcohol control policies (Ferreira-Borges et al., 2017; Morojele et al., 2021). In contrast, NAFLD, while currently under-recognized and less prevalent, is emerging as a significant health concern in several middle-SDI countries, driven by increasing obesity, type 2 diabetes, urbanization, and nutritional transitions (Spearman et al., 2021). Its growing impact signals a shifting liver disease landscape and raises questions about health system readiness for non-communicable conditions that demand different models of care (Beaglehole et al., 2008; Spearman et al., 2021; Younossi et al., 2016). The inverse association between SDI and HBV-related burden, alongside rising NAFLD in more developed settings, exemplifies a broader epidemiological transition also observed in other low- and middle-income regions (Beaglehole et al., 2008; Golabi et al., 2021; Spearman et al., 2021). These shifts highlight the urgent need for tailored, dual-pronged strategies: sustaining viral hepatitis elimination efforts while preparing systems to detect and manage metabolic liver disease (Younossi et al., 2024). Failing to act on both fronts risks further exacerbating liver disease burdens in already strained health systems across SSA.

Looking ahead, projections through 2035 suggest that while some gains are likely, cirrhosis and chronic liver disease will remain a significant and unevenly distributed health challenge across SSA. Overall ASDRs and DALYs are expected to decline modestly across the region, but substantial geographic variation will persist, with Central and Eastern SSA continuing to carry the highest burden (Xiao et al., 2023). Notably, our projections indicate that males will consistently experience nearly twice the burden of cirrhosis-related mortality and morbidity compared to females—a pattern that shows little sign of narrowing over time (Jepsen & Younossi, 2021; Tan et al., 2023). These persistent disparities highlight systemic gaps in early detection, gender-sensitive care, and equitable service delivery (Achrekar et al., 2024; Yaya et al., 2020). Encouragingly, modelled interventions such as expanded hepatitis B vaccination illustrate the tangible benefits of prevention, with a projected 27% reduction in HBV-related mortality under a scale-up scenario. However, such gains are unlikely to materialize without dedicated investment in programmatic delivery, surveillance systems, and region-specific strategies (Yaya et al., 2020). Critically, health systems across SSA remain underprepared to adapt to shifting patterns of liver disease—whether through expanding primary care capacity, integrating liver health into universal health coverage agendas, or strengthening long-term care models (Atun et al., 2017; Boudreaux et al., 2020). Future progress will require not only sustaining momentum on infectious disease control, but also reorienting health policy to anticipate and address evolving demographic and epidemiological pressures. Without timely and context-specific interventions, projected improvements may plateau, leaving the most vulnerable populations behind.

## Strengths and Limitations

This study provides the most comprehensive assessment to date of liver cirrhosis and chronic liver diseases across SSA by leveraging the extensive GBD 2021 data. Its detailed aetiologic stratification—including hepatitis B, hepatitis C, alcohol-related liver disease, and NAFLD— and use of SDI offer nuanced insights into geographic, temporal, and sex-specific patterns. Projected future burdens under different scenarios further inform policy and health system planning amid epidemiological transitions. However, limitations include reliance on secondary data of variable quality and completeness across countries, especially in low-SDI and conflict-affected regions, which may lead to underreporting or misclassification. Attribution of burden to specific causes is challenging due to overlapping risk factors and diagnostic constraints, potentially biasing estimates. Projections, while robust, cannot fully anticipate future changes in healthcare, behaviour, or interventions. The ecological nature of the data also restricts exploration of individual-level risk factors, limiting precision in targeting interventions. Finally, the current low prevalence of NAFLD likely underrepresents its growing impact, emphasizing the need for improved surveillance. Despite these limitations, the study’s broad geographic and aetiologic coverage, sex and socio-demographic analyses, and forward-looking approach provide essential evidence to guide integrated strategies addressing infectious and metabolic liver disease burdens in SSA.

## Conclusion

Our findings reveal that despite measurable progress in reducing cirrhosis-related mortality rates across SSA, the growing absolute burden and persistent disparities by geography, sex, and socioeconomic status pose significant challenges. This evolving landscape demands urgent, context-specific health system reforms that integrate robust viral hepatitis control with emerging strategies to address metabolic liver disease. Achieving equitable liver health outcomes will depend on strengthening prevention, early diagnosis, and treatment programs— especially in underserved and high-risk populations—while anticipating demographic and epidemiological shifts. Ultimately, addressing the dual burden of infectious and non-communicable liver diseases in SSA is critical to curbing avoidable morbidity and mortality and advancing regional health equity.

## Acknowledgements

This work was inspired by the 2025 King’s Global Engagement Partnership award, which facilitated new collaborations in sub-Saharan Africa, and was conducted in partnership with students through the King’s Undergraduate Research Fellowship (KURF). We gratefully acknowledge these initiatives at King’s College London for fostering an environment that supports impactful and globally relevant research. We also thank colleagues and collaborators who continue to contribute valuable insights to this work.

## Author Contributions

E.K.P. and D.L.S.: conceptualization; E.K.P., K.Y., and R.L.: data curation; E.K.P.: data analysis; D.L.S.: supervision; D.L.S. and E.K.P.: validation; E.K.P.: writing—original draft; all authors (E.K.P., R.L., K.Y., and D.L.S.) contributed to writing—review and editing, and all authors read and approved the final version to be submitted.

## Data Availability Statement

All data generated or analysed during this study are included in this published article and its Supplementary Material. Further inquiries can be directed to the corresponding author.

