## Supplementary File 1 for "Trends and Future Projections of Liver Cirrhosis Burden in Sub-Saharan Africa with Hepatitis B Vaccination Impact"

**Table 1: Geographic Classification of Sub-Saharan Africa According to the Global Burden of Disease (GBD) Database**

| **Super region** | **4 sub regions** | **Countries and territories** |
|---|---|---|
| Sub-Saharan Africa | Central sub-Saharan Africa | Angola, Central African Republic, Congo (Brazzaville), DR Congo, Equatorial Guinea, Gabon |
| | Eastern sub-Saharan Africa | Burundi, Comoros, Djibouti, Eritrea, Ethiopia, Kenya, Madagascar, Malawi, Mozambique, Rwanda, Somalia, South Sudan, Uganda, Tanzania, Zambia |
| | Southern sub-Saharan Africa | Botswana, Eswatini, Lesotho, Namibia, South Africa, Zimbabwe |
| | Western sub-Saharan Africa | Benin, Burkina Faso, Cabo Verde, Cameroon, Chad, Côte d'Ivoire, The Gambia, Ghana, Guinea, Guinea-Bissau, Liberia, Mali, Mauritania, Niger, Nigeria, São Tomé and Príncipe, Senegal, Sierra Leone, Togo |

**Table 2: Socio-Demographic Index (SDI) Values and Quintile Classification for Sub-Saharan Africa Locations, GBD 2021**

| Socio-demographic Index values for all estimated GBD 2021 SSA locations | | |
|---|---|---|
| **Location** | **SDI values** | **SDI quintile** |
| **Sub-Saharan Africa** | 0.458587301 | Low SDI |
| **Central sub-Saharan Africa** | 0.472255651 | Low-middle SDI |
| Angola | 0.453721949 | Low SDI |
| Central African Republic | 0.30916769 | Low SDI |
| Congo (Brazzaville) | 0.583075236 | Low-middle SDI |
| DR Congo | 0.383179849 | Low SDI |
| Equatorial Guinea | 0.657857456 | Middle SDI |
| Gabon | 0.634691393 | Middle SDI |
| **Eastern sub-Saharan Africa** | 0.409720983 | Low SDI |
| Burundi | 0.289374365 | Low SDI |
| Comoros | 0.475978688 | Low-middle SDI |
| Djibouti | 0.487958371 | Low-middle SDI |
| Eritrea | 0.403863943 | Low SDI |
| Ethiopia | 0.358823295 | Low SDI |
| Kenya | 0.523768077 | Low-middle SDI |
| Madagascar | 0.400246943 | Low SDI |
| Malawi | 0.384553634 | Low SDI |
| Mozambique | 0.326462614 | Low SDI |
| Rwanda | 0.435588706 | Low SDI |
| Somalia | 0.077688109 | Low SDI |
| South Sudan | 0.278371125 | Low SDI |
| Uganda | 0.423261181 | Low SDI |
| Tanzania | 0.446568273 | Low SDI |
| Zambia | 0.505948954 | Low-middle SDI |
| **Southern sub-Saharan Africa** | 0.642200282 | Middle SDI |
| Botswana | 0.642721629 | Middle SDI |
| Eswatini | 0.585459713 | Low-middle SDI |
| Lesotho | 0.510393066 | Low-middle SDI |
| Namibia | 0.617564872 | Low-middle SDI |
| South Africa | 0.679626598 | Middle SDI |
| **Western sub-Saharan Africa** | 0.446022979 | Low SDI |
| Benin | 0.373486574 | Low SDI |
| Burkina Faso | 0.285118402 | Low SDI |
| Cabo Verde | 0.533534539 | Low-middle SDI |
| Cameroon | 0.479691223 | Low-middle SDI |
| Chad | 0.240436019 | Low SDI |
| Côte d'Ivoire | 0.425941883 | Low SDI |
| The Gambia | 0.40971416 | Low SDI |
| Ghana | 0.56493039 | Low-middle SDI |
| Guinea | 0.336401293 | Low SDI |
| Guinea-Bissau | 0.353109621 | Low SDI |
| Liberia | 0.352442452 | Low SDI |
| Mali | 0.268579941 | Low SDI |
| Mauritania | 0.4989451 | Low-middle SDI |
| Niger | 0.168072774 | Low SDI |
| Nigeria | 0.503390833 | Low-middle SDI |
| São Tomé and Príncipe | 0.505413747 | Low-middle SDI |
| Senegal | 0.408054193 | Low SDI |
| Sierra Leone | 0.358665881 | Low SDI |
| Togo | 0.408533695 | Low SDI |

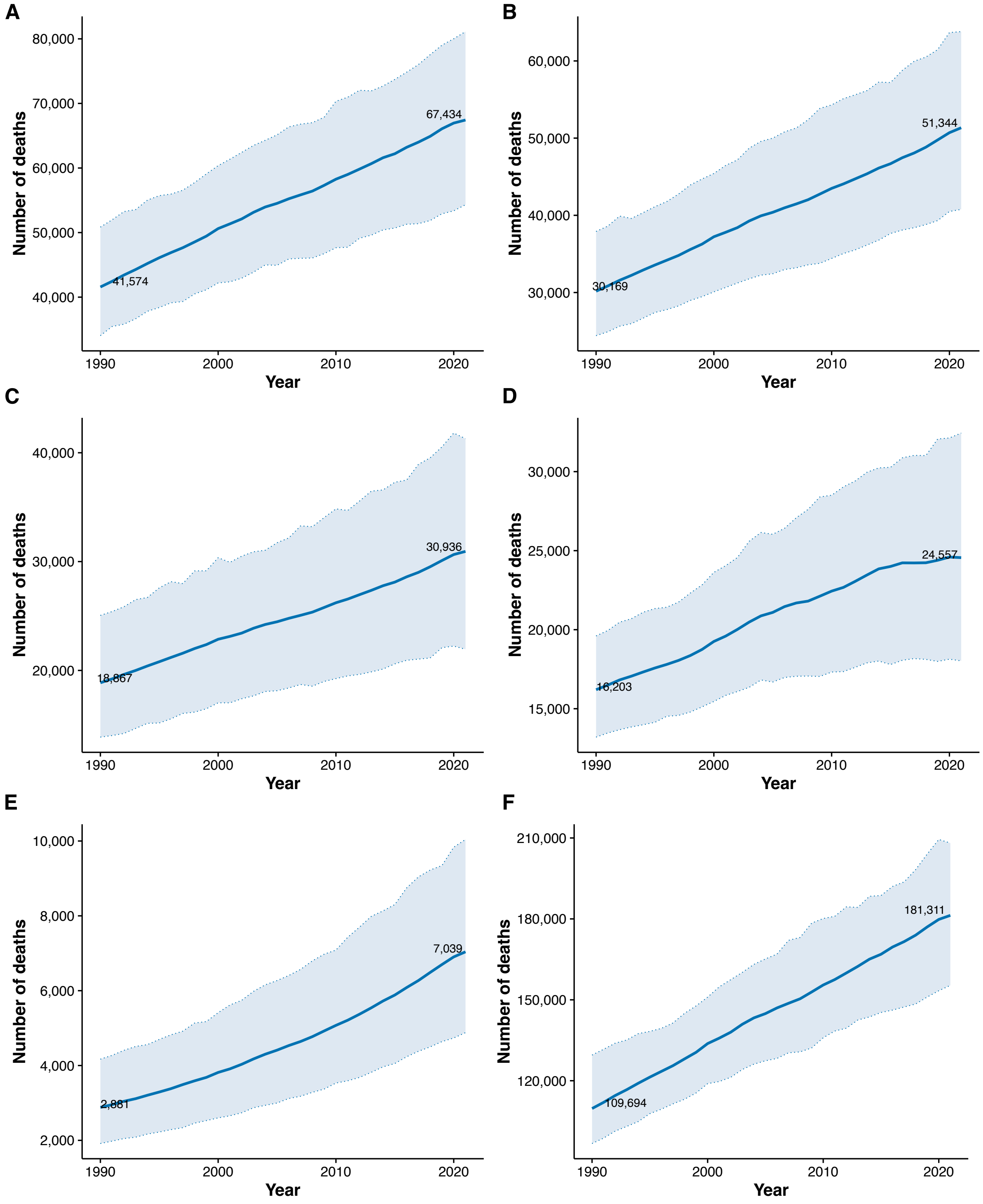

**Supplementary Figure 1. Trends in total and cause-specific liver cirrhosis deaths in Sub-Saharan Africa, 1990–2021**. Temporal trends in cirrhosis deaths attributable to hepatitis B virus (A), hepatitis C virus (B), alcohol use (C), non-alcoholic fatty liver disease (D), other causes (E), and total cirrhosis deaths (F). Lines represent mean estimates; shaded ribbons indicate 95% uncertainty intervals.

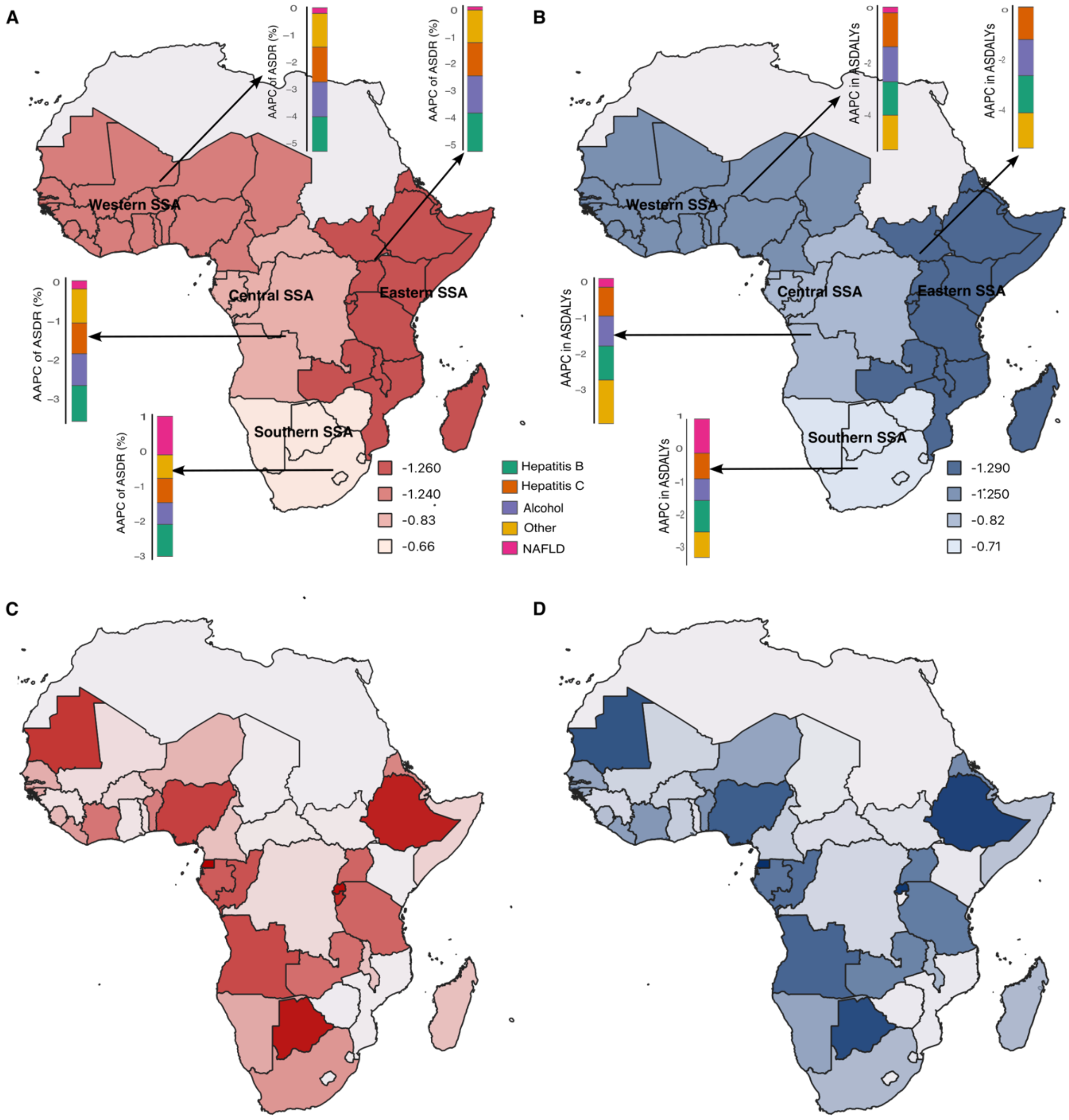

**Supplementary Figure 2. Average annual percent change (AAPC) for cirrhosis aetiologies and cirrhosis burden across Sub-Saharan Africa, 1990–2021.** AAPC in ASDR (A) and ASDALYs (B) by sub-region. Bars for each sub-region represent AAPC estimates by cirrhosis aetiology—hepatitis B, hepatitis C, alcohol use, non-alcoholic fatty liver disease, and other causes. AAPC in ASDR (C) and ASDALYs (D) at the country level across SSA from 1990–2021. Deeper red or blue colours indicate more negative AAPCs, reflecting faster declines in cirrhosis burden.

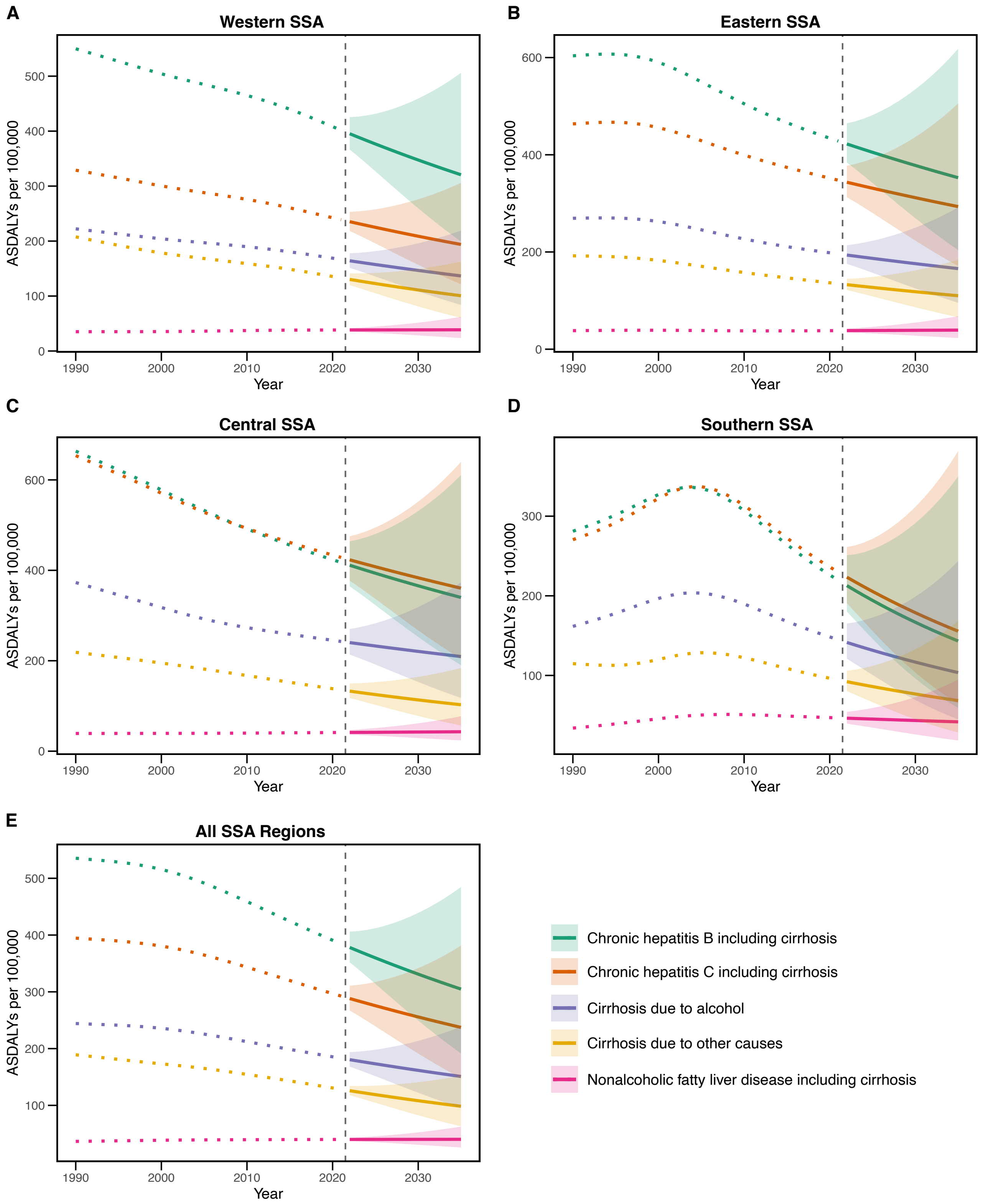

**Supplementary Figure 3. Projected disability-adjusted life years (ASDALYs) for cirrhosis aetiologies across Sub-Saharan Africa, 2022–2035.** Panels (A)–(E) show ASDALY projections per 100,000 population for Western SSA, Eastern SSA, Central SSA, Southern SSA, and total SSA, respectively. Coloured dotted lines represent observed data for each aetiology from 1990 to 2021, while solid lines indicate corresponding projections from 2022 to 2035. Shaded ribbons denote 95% uncertainty intervals around the projections. The vertical dashed line marks the boundary between observed and projected periods.

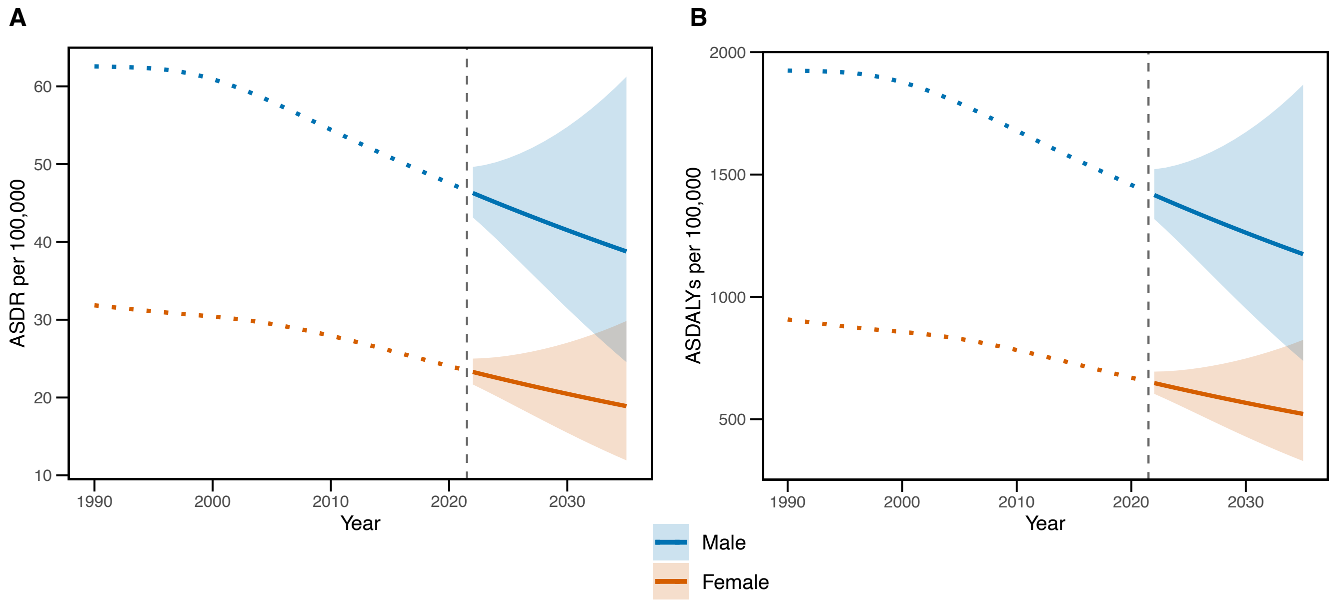

**Supplementary Figure 4. Projected ASDR and ASDALYs by sex for cirrhosis and other chronic liver diseases across Sub-Saharan Africa, 2022–2035.** Panels show ASDRs (A) and ASDALYs (B) per 100,000 population for the whole SSA region. Coloured dotted lines represent observed data from 1990 to 2021, while solid lines indicate projections from 2022 to 2035. Shaded ribbons denote 95% uncertainty intervals. The vertical dashed line marks the transition from observed to projected data.
